# Resolving the diagnostic odyssey in inherited retinal dystrophies through long-read genome sequencing

**DOI:** 10.1101/2024.08.28.24312668

**Authors:** Gerardo Fabian-Morales, Vianey Ordoñez-Labastida, William J. Rowell, Christine Lambert, Cairbre Fanslow, Alexander Robertson, Juan C. Zenteno

**Affiliations:** Department of Genetics, Institute of Ophthalmology "Conde de Valenciana", Mexico City, Mexico; Rare Disease Diagnostic Unit, Faculty of Medicine, UNAM, Mexico City, Mexico; Faculty of Medicine, Autonomous University of the State of Morelos (UAEM), Morelos, Mexico; PacBio, Menlo Park, CA, USA; Faculty of Medicine, Department of Biochemistry, National Autonomous University of Mexico (UNAM), Mexico City, Mexico

**Keywords:** Retinal dystrophy, long reads-genome sequencing, intronic variant, structural variants

## Abstract

**Background:** Inherited Retinal Dystrophies (IRDs) are visually disabling monogenic diseases with remarkable genetic and phenotypic heterogeneity. Mutations in more than 300 different genes have been identified as disease causing. Genetic diagnosis of IRDs has been greatly improved thanks to the incorporation of Next Generation Sequencing (NGS) approaches. However, the current IRD molecular diagnosis yield using NGS is approximately 60% and negative cases can be explained by variants that are not usually identified by the widely used short reads-NGS such as structural variants (SVs) or by variants located in uncovered, low complexity, repetitive, highly homologous, or GC-rich regions. Long-read genome sequencing (LR-GS) is an emerging technology that produces 10-20 kb reads and is expected to overcome short-read sequencing limitations in the clinical context, thus improving the diagnostic yield in heterogeneous diseases as IRDs.

**Objective:** To describe LR-GS utility in 3 unrelated, previously unsolved IRD cases.

**Material & Methods:** LR-GS was performed on 3 probands with IRDs and previous inconclusive genetic testing with NGS (either exome or gene panel sequencing). Whole genome libraries were prepared using SMRTbell® prep kit. Sequencing was performed on the PacBio Revio system.

**Results:** A definite diagnosis was established in the 3 cases. A homozygous deep intronic variant c.4885+740A>T in *USH2A* was identified in a proband with Usher syndrome; A homozygous intragenic deletion involving *EYS* exon 24 was found in a proband with Retinitis pigmentosa. Finally, a proband with Usher syndrome was found to be a compound heterozygous for a *USH2A* deep intronic variant and a multiexonic duplication involving *USH2A* exons 22-32.

**Conclusion:** Our case series show the efficiency in a clinical setting of LR-GS to detect disease-causing variants that were missed by current NGS techniques, improving thus the molecular diagnosis rate in genetically heterogeneous diseases as IRDs

## INTRODUCTION

Inherited retinal dystrophies (IRDs) are monogenic inherited conditions characterized by severe visual impairment due to progressive loss of cells from the retina and retinal pigment epithelium (Murro et al., 2023). IRDs are one of the most genetically heterogeneous diseases in humans as mutations in approximately 300 different genes are known to underlie a diversity of retinal dystrophic phenotypes (RetNet, accessed in July 2024). The incorporation of Next Generation Sequencing technologies (NGS) such as exome sequencing (ES) or genome sequencing (GS) in clinical settings, has enabled efficient genetic diagnosis in IRD patients, greatly broadening the knowledge of the genetic landscape leading to inherited retinal blindness (Britten-Jones et al., 2023). Nevertheless, the current IRD diagnosis yield using NGS usually ranges between 50% and 70% (Carss et al., 2017; Dockery et al., 2021, Britten-Jones et al., 2022; De Brujin et al., 2023; Weisschuh et al., 2024). Among other reasons, negative cases could be explained either by: 1) mutations in genes that have not been previously associated with IRDs; 2) sequencing limitations e.g., GC-rich regions; or, 3) alignment limitations including low complexity, repetitive regions, segmental duplications, or variants that are not usually detected by short-read (SR) based NGS such as structural Variants (SVs) (Bruijn et al., 2021; Olivucci et al., 2024).

Third generation sequencing, also known as Long Read Sequencing (LRS) is a PCR- free and real-time sequencing process that produces 10-20 kb reads on average and is expected to overcome short-read sequencing limitations (Olivucci et al., 2024), thus improving the diagnostic yield in genetically heterogeneous diseases. Although LRS is not currently widely implemented in the clinical context, there is an increasing number of studies that show the value of this technology for diagnosis of genetic conditions like IRD, especially in cases unsolved after ES or SR GS (Sano et al., 2022; Li et al., 2024).

The benefits of LRS in clinical settings encompass, among others, the detection of balanced and unbalanced SVs including complex rearrangements and their precise breakpoint definition and the identification of deep intronic variants and expansion repeats in a single assay (Olivucci et al., 2024). Targeted LRS has also proved its utility in IRD diagnosis by detecting SNVs and indels with high accuracy in low complexity regions that are not effectively covered by SR sequencing, such as *RPGR* ORF15 (Bonetti et al., 2023).

In this work, we describe the employ of LR-genome sequencing (LR-GS) to genetically solve three unrelated IRD cases who had negative results after SR GS. Our results stress the potential utility of LR-GS for the identification of elusive disease-causing variants and for the improvement of diagnostic rates in genetically heterogeneous disorders as IRDs.

## Material and Methods

The study was approved by the Institutional Review Board of the Institute of Ophthalmology “Conde de Valenciana’’ (Mexico City, Mexico) (CEI-2021/12/01). Written informed consent was obtained from all subjects or their parents. The study was conducted in accordance with the Declaration of Helsinki.

### LR-GS

#### Library Preparation and Sequencing

Samples were prepared following the standard operating procedure (SOP) available at PACB.com ("Preparing whole genome and metagenome libraries using SMRTbell® prep kit 3.0"). Sequencing was performed on the PacBio Revio system with the Revio Polymerase Kit, following the Revio SMRT Link setup. Genomic DNA quality and concentration were assessed using the FEMTO Pulse (Agilent Technologies) and Qubit dsDNA HS reagents Assay kit (Thermo Fisher Scientific). SMRTbell libraries were constructed on the Hamilton Microlab Star and VENUS 5 system following the SOP available on PACB.com under "Preparing whole genome and metagenome libraries using SMRTbell® prep kit 3.0". Completed libraries were size-selected for fragments larger than 10 kb using the PippinHT system with "6-10 kb High Pass Marker 75E" cassette definition (Sage Science). For library characterization, post size-selected SMRTbell Library samples were quantified using the Qubit DNA HS assay and DNA size was estimated using the FEMTO Pulse. The libraries were loaded at an On-Plate Concentration of 90 pM.

### Bioinformatic analysis

The raw sequencing data were processed with the PacBio WGS Pipeline HiFi-human-WGS-WDL (v1.0.3). Sequencing reads were aligned to the GRCh38/Hg38 using pbmm2 (v1.10.0) with default parameters. DeepVariant (v1.5.0) was used for SNPs and small indels calling while PBSV (v2.9.0) was applied for SVs calling and HiFiCNV (v0.1.7) for advanced CNV calling. Small variant annotation was performed with slivar (v0.2.2) and bcftools (1.14) and structural variant annotation using svpack. Franklin Genoox (https://franklin.genoox.com/clinical-db/home) and Geneyx (https://geneyx.com/) tools were used for variant interpretation and prioritization.

### Patients

This is a serie of 3 IRDs cases, 2 females and 1 male. Patients were 38 years old on average at the moment of the study.

#### Case #1

This patient was born from a consanguineous couple arising from a small village and who complained of night blindness, progressive visual loss, and hypoacusia since middle childhood. A clinical diagnosis of Usher syndrome was provided more than 10 years ago. Noteworthy, this proband has a sibling of similar age affected with the same disorder. Previous genetic screening in the proband included sequencing of a gene panel containing 293 genes related to retinal dystrophies (Invitae, San Francisco CA) which failed to identify pathogenic SNVs/ CNVs.

#### Case #2

This patient has a history of night blindness and progressive visual loss since early adulthood. There is no parental consanguinity. The proband has a younger sibling with progressive visual loss of unknown cause. A clinical diagnosis of retinitis pigmentosa (RP) was established in middle adulthood. Previous genetic screening included exome sequencing (3billion, Seoul, Korea) but no causal SNVs or CNVs were recognized.

#### Case #3

This proband has a history of decreased visual acuity and hearing loss since early adulthood. There was no parental consanguinity nor additional affected relatives in the family. Previous genetic screening included sequencing of a panel of 330 retinal dystrophy genes (Invitae) which disclosed the presence of both a pathogenic heterozygous duplication of *USH2A* exons 22-32 and a heterozygous *USH2A* c.11366T>C (p.Ile3789Thr) classified as VUS and with an extremely low frequency in GnomAD (40 alleles out of 1,613,604) with no instances of homozygosity. However, subsequent Sanger sequencing and qPCR assays in DNA from several members of the family disclosed that both the duplication and the point variant localize in the same *USH2A* allele (*cis*) and thus, the case was considered not genetically solved. Subsequently, exome sequencing was performed in the proband’s DNA, without the identification of causative variants.

### LR-GS results

#### Case #1

In this proband with clinical diagnosis of Usher syndrome, homozygosity for a deep intronic c.4885+740A>T variant in *USH2A* intron 23 was identified (Figure #1). Splice AI predicts this variant as splice-altering with a 0.7 (strong) score. This variant is absent from GnomAD (Genome), is extremely rare in TOPMed Bravo (6 Alleles out of 264,690) and is present in 15 /19,896 alleles from the Mexico City Prospective Study (MCPS, https://rgc-mcps.regeneron.com/rsid/rs960250478). Subsequent direct Sanger sequencing in DNA from the affected sibling identified homozygosity for the novel deep intronic variant in *USH2A*. Parental DNA samples were not available for analysis.

**Figure 1.**
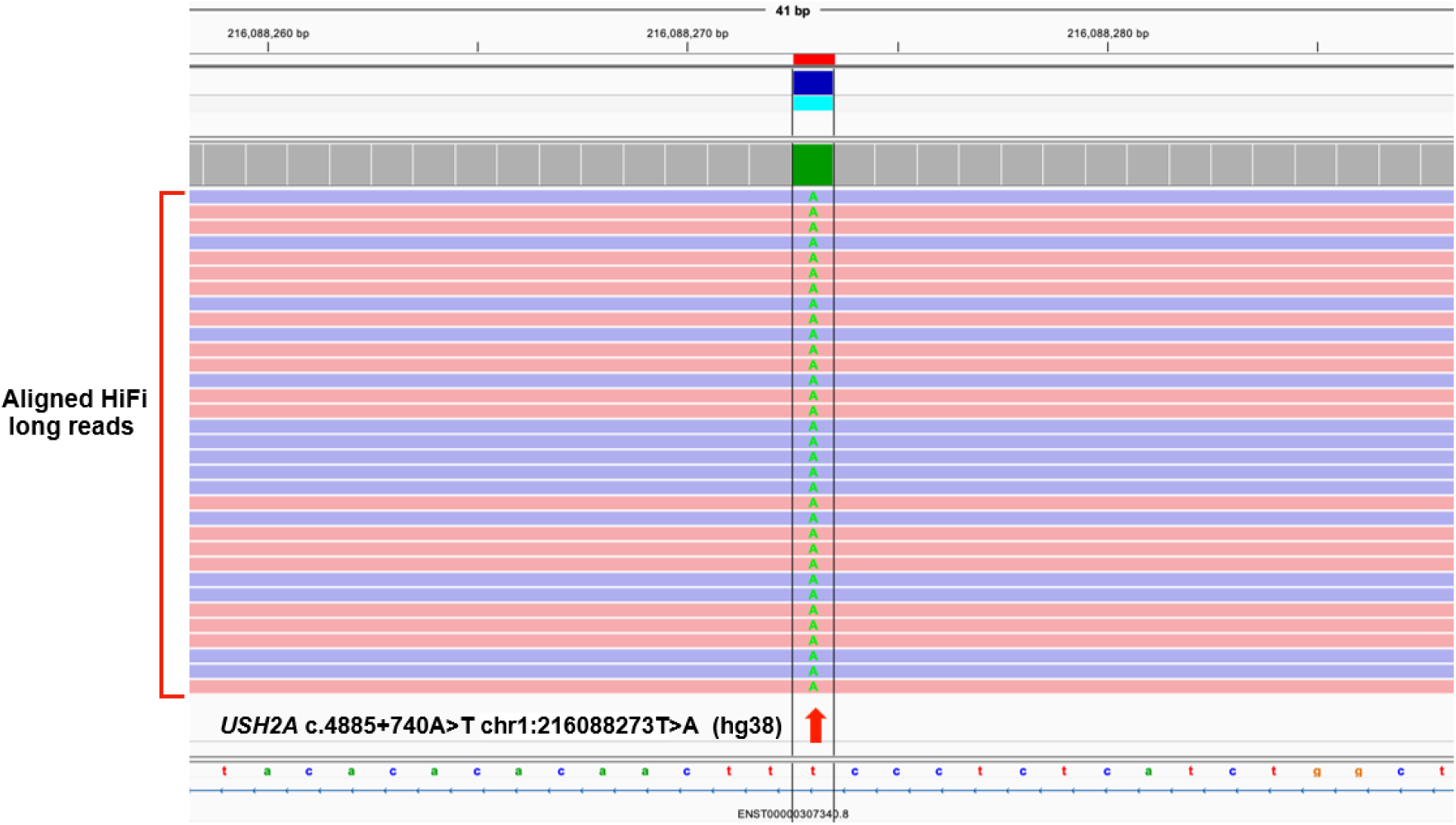
Visualization of patient #1 long reads in IGV showing homozygosity for the deep intronic c.4885+740A>T chr1:216088273T>A (hg38) in *USH2A*.

#### Case #2

In this proband with a diagnosis of retinitis pigmentosa, LRS allowed the recognition of a homozygous deletion of exon 24 of the *EYS* gene. The deletion spanned 13 kb (DEL:chr6:64605825-64618845, hg38 reference genome) (Figure #2), has been previously identified in compound heterozygous state with a pathogenic SNV in a patient with retinitis pigmentosa (unpublished results) and has been reported once in ClinVar (accession number 644814).

**Figure 2.**
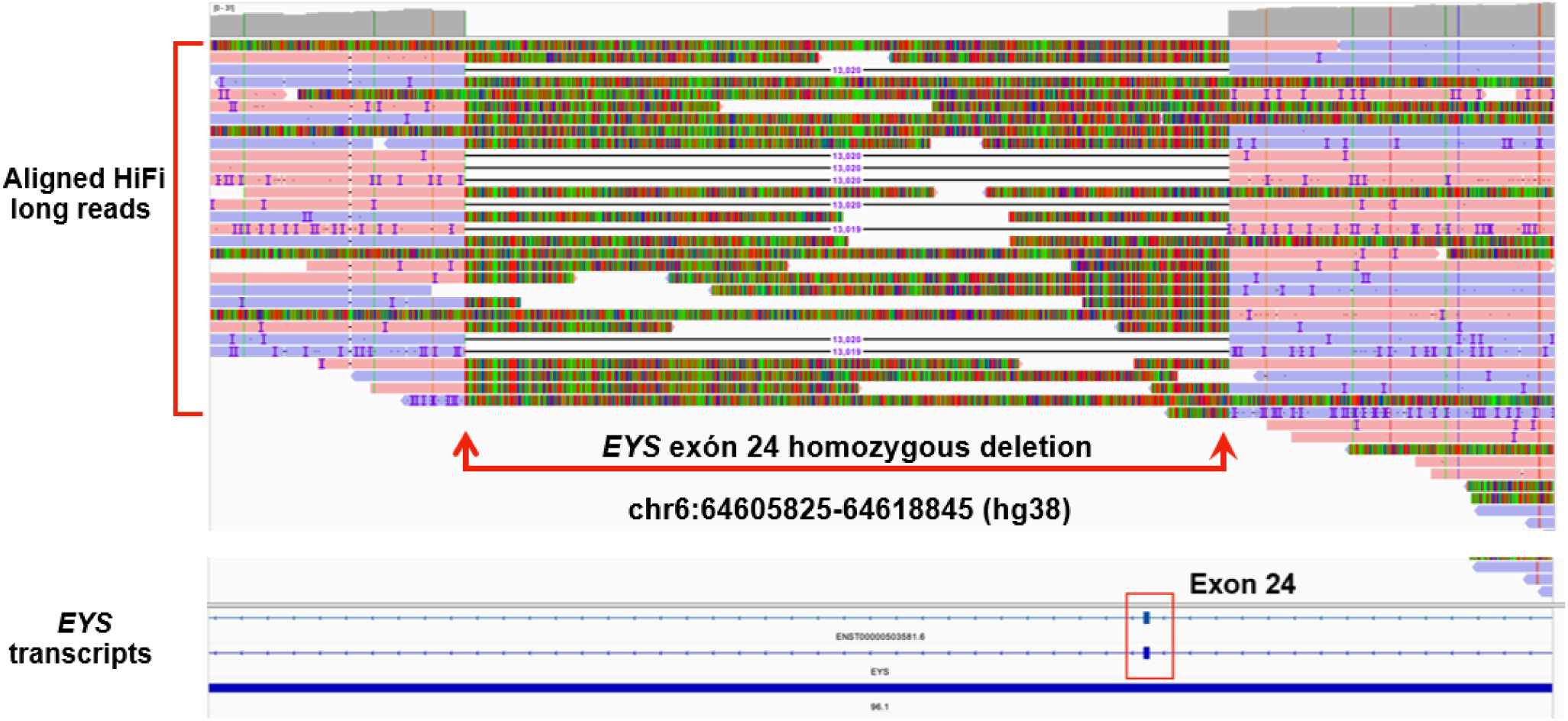
Visualization of patient #2 long reads in IGV indicate a homozygous 13 kb intragenic deletion in *EYS* gene at 6q12 (chr6:64605825-64618845, hg38). A complete drop in the coverage within the breakpoints (red arrows) due to the absence of alignments is observed.

#### Case #3

In this patient with a clinical diagnosis of Usher syndrome, LRS confirmed a duplication of exons 22-32 of *USH2A* (DUP:chr1:216023829-216099513, hg38 reference genome) previously identified by a gene panel sequencing. In addition, LRS allowed for phasing of this CNV and a concurrent heterozygous c.11366T>C (p.Ile3789Thr) *USH2A* variant of unknown significance and confirmed that they reside in *cis*. Subsequent deep visual analysis of the entire *USH2A* locus resulted in the identification of a c.4885+740A>T variant in *USH2A* intron 23, which is predicted to have a strong splice-altering effect and is absent from public databases, except TOPMed (6 out of 264,690 alleles) and MCPS (15 out of 19,896 alleles). Of note, as the intronic variant was located within the duplicated region in the other *USH2A* allele, the allele balance of the splice variant was affected in long reads results (Figure #3). Finally, segregation analysis by Sanger sequencing confirmed that the intronic *USH2A* variant was inherited from one parent (supplementary figure) while qPCR assays demonstrated that the other parent was carrier of the *USH2A* exons 22-32 duplication.

**Figure 3.**
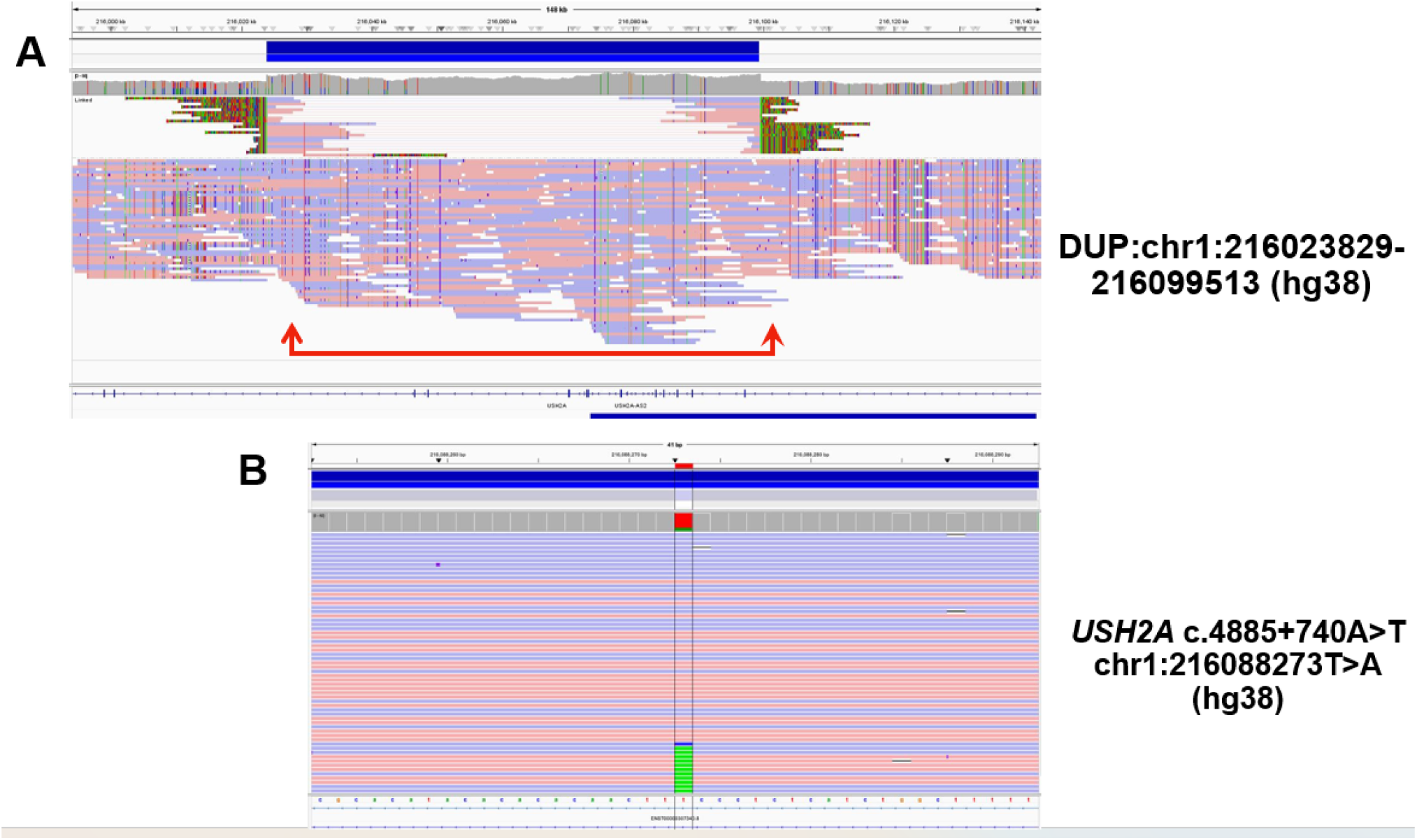
Visualization of patient #3 long reads in IGV. (A) A heterozygous 75.6 kb duplication at 1q41 (chr1:216023829-216099513; hg38) encompassing *USH2A* exons 22-32 is indicated by increased number of reads (red arrows). (B) Green rectangles indicate reads with the deep intronic c.4885+740A>T, chr1:216088273T>A (hg38) in *USH2A.* Note that the intronic variant is located within the duplicated region in the other *USH2A* allele causing its underrepresentation in sequencing reads.

## DISCUSSION

Although NGS has revolutionized the process of genetic diagnosis of human diseases, various technical limitations need to be overcome for its optimal implementation. For example, SR-based NGS usually produces sequence reads of 100 or 150 bp in length that are very efficient in detecting simple nucleotide substitutions and indels but can overlook extensive genomic rearrangements, including retrotransposon insertions, repeat expansions, and copy number neutral inversions (Mantere et al., 2019). In contrast, LRS provides sequence reads ranging between 10 kb and 100 kb and consequently is capable of identifying complex structural variants (Conlin et al., 2022). In addition, LRS avoids biases associated with PCR amplification since isolated DNA can be used directly for sequencing without the need of cluster generation.

Recent advances in LRS have enabled researchers to identify several previously unrecognized genetic variants (Reviewed in: Mastrorosa et al., 2023; Olivucci et al., 2024). Particularly, LRS has been successfully employed for molecular characterization of the genetically heterogeneous IRDs. For example, LR-GS was able to successfully identify multi-exonic deletions (Sano et al., 2022), deep intronic mutations (Li et al, 2024), and inverted duplications (Fadaie et al., 2021). In another recent example using targeted LRS, a pathogenic heterozygous Alu element insertion missed by SRS was identified (Fernandez-Suárez et al., 2024).

In this work, we present a case series of 3 unrelated patients with inherited retinal dystrophies in whom previous NGS using SR sequences was unable to identify the causal genetic defects. A total of 6 pathogenic variants, including a novel recurrent deep intronic variant in *USH2A* (3 alleles), a homozygous single exon deletion in *EYS* (2 alleles) and a multi-exon duplication in *USH2A* (1 allele), were characterized.

The novel *USH2A* c.4885+740A>T deep intronic variant in intron 23 was identified as homozygous in a pair of Usher syndrome siblings and as heterozygous in a sporadic Usher syndrome case (patient #3) carrying a pathogenic *USH2A* intragenic duplication in *trans*. The criteria to consider this deep intronic variant as disease-causing includes: 1) *in silico* splice-altering prediction with a donor gain delta score of 0.7 (2 bp) and acceptor gain delta score of 0.43 (155bp) in Splice AI, and a splice gain score of 0.43 in Pangolin; 2) its absence in public databases; and 3) its segregation with the disease in affected cases. Interestingly, directed Sanger analysis of additional syndromic and non-syndromic IRD cases with a single pathogenic variant in *USH2A* from our Institution demonstrated that 6 individuals carried the c.4885+740A>T deep intronic *USH2A* variant, which allowed the genetic resolution of such cases (data not shown). Remarkably, all 8 subjects arose from the same region, suggesting a founder mutation effect. This variant is absent from public databases but is included at low frequency in the MCPS database where it was identified exclusively in individuals with indigenous background, supporting a founder mutation effect. In addition, in patient #3, LRS successfully determined the *trans* configuration of *USH2A* variants by phasing causal mutations in a single assay. Homozygous or compound heterozygous mutations in the *USH2A* gene, located in 1q41, are the cause of 12%–25% of autosomal recessive Retinitis Pigmentosa cases (McGee et al., 2012; Verbakel et al., 2018). *USH2A* is a large gene composed of 72 exons and its mutational spectrum includes approximately 1,700 variants classified as pathogenic or likely pathogenic by the ACMG (LOVD and HGMD databases, accessed in July 2024). Disease-causing mutations include missense, nonsense, deletions, duplications, and splicing variants (Su et al., 2022). With the advent of WGS, several pathogenic deep intronic variants have been identified (Reurink et al., 2023). It is probable that novel SVs and deep-intronic variants that cause aberrant mRNA splicing will be increasingly recognized when genetically unsolved cases are analyzed with WGS, as the cases presented here. Nonetheless, functional analyses as minigene assays will be needed to confirm the pathogenicity of the novel c.4885+740A>T variant in *USH2A* identified here.

In another patient, LRS efficiently identified a homozygous deletion of *EYS* exon 24 which was missed in a previous SRS exome analysis. *EYS* variants account for 16-23% of autosomal recessive RP cases in diverse populations (Barragán et al., 2010; Yang et al. 2020). IRD-causative CNVs in *EYS* are relatively common and are particularly located in exons 12–22 (Xu et al., 2021). One study found that pathogenic CNVs in *EYS* constitute the second pathogenic allele in ∼15% of cases where a mutation was previously detected by direct sequencing (Pieras et al., 2011). The exon 24 deletion identified here is included in ClinVar (accession number 644814) and has been recognized by our group *in trans* to a pathogenic missense variant in a IRD patient from the same country (unpublished data).

In conclusion, our case series demonstrates the power of LRS for genetic resolution of IRD cases which were molecularly uncharacterized using short read NGS techniques. LRS will significantly increase the diagnostic yield for patients in which the causative genetic defect remains elusive after traditional NGS approaches.

## Data Availability

All data produced in the present work are contained in the manuscript

---

**Supplementary figure.**
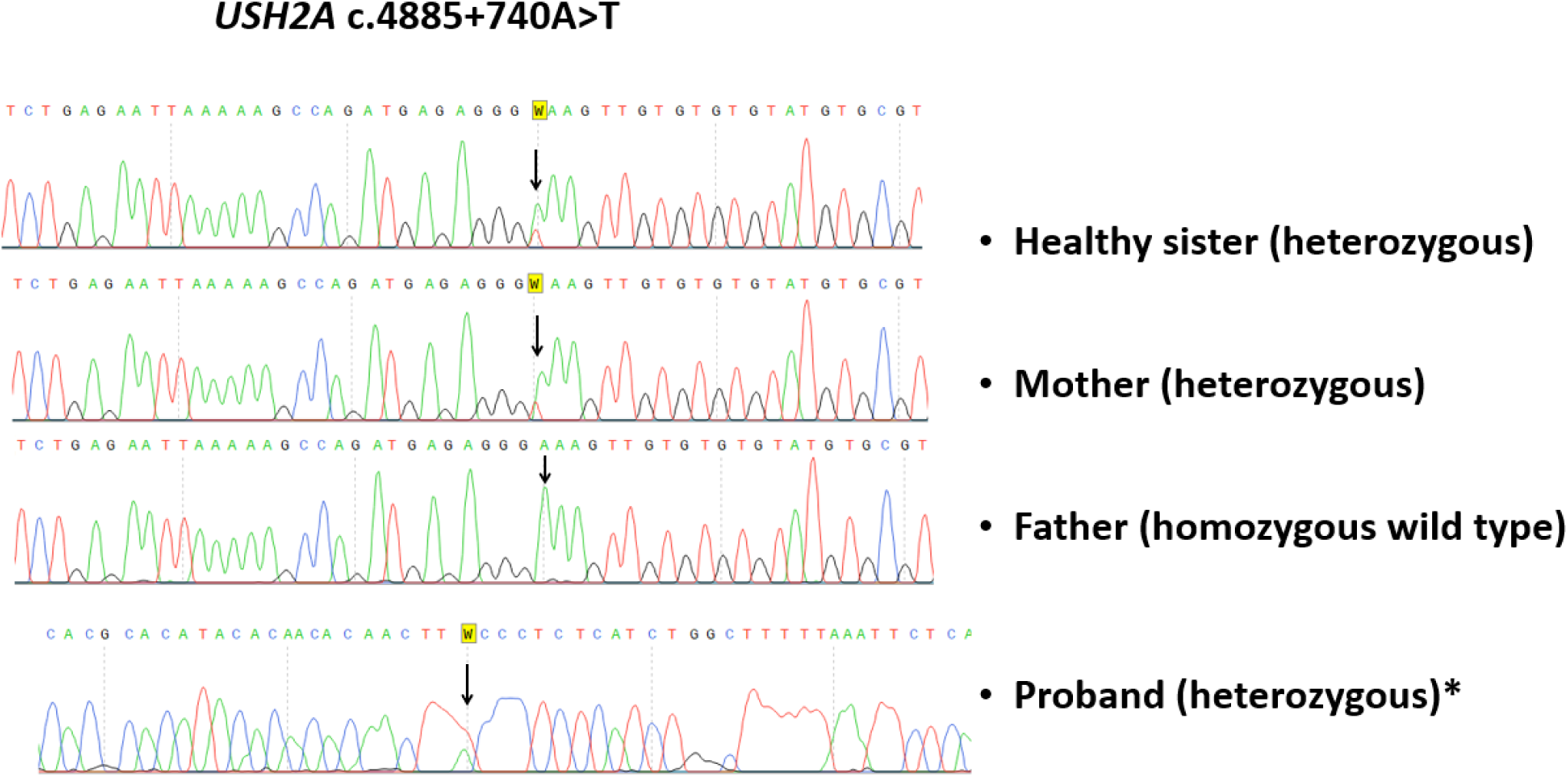
Sanger sequencing analysis of the novel *USH2A* c.4885+740A>T (chr1:216088273T>A) deep intronic variant in DNA from patient #3 and his relatives.

## Notes

### Competing Interest Statement

The authors have declared no competing interest.

### Funding Statement

This study did not receive any funding

### Author Declarations

Institutional Review Board of The Institute of Ophthalmology 'Conde de Valenciana' gave ethical approval for this work

